# Missed opportunities of flu vaccination in Italian target categories: insights from the online EPICOVID 19 survey

**DOI:** 10.1101/2020.09.30.20204560

**Authors:** Andrea Giacomelli, Massimo Galli, Stefania Maggi, Gabriele Pagani, Raffaele Antonelli Incalzi, Claudio Pedone, Mauro di Bari, Marianna Noale, Caterina Trevisan, Fabrizio Bianchi, Marcello Tavio, Massimo Andreoni, Claudio Mastroianni, Aleksandra Sojic, Federica Prinelli, Fulvio Adorni, on Behalf of EPICOVID19 Working Group

## Abstract

We aimed to assess the reported rate of flu vaccination in the season 2019/2020 in respondents to the Italian nationwide online EPICOVID 19 survey. A national convenience sample of volunteers aged 18 or older was assessed between 13^th^ April and 2^nd^ June 2020. Flu vaccine rates were calculated for all classes of age. The association between the independent variables and the flu vaccine was assessed by applying a multivariable binary logistic regression model. Of the 198,822 respondents 41,818 (21.0%) reported to have received a flu vaccination shot during the last influenza season. In particular, 15,009 (53.4%) subjects aged 65 years or older received a flu vaccination shot. Being 65 years aged or older (aOR 3.06, 95%CI 2.92-3.20) and having a high education level (aOR 1.34. 95%CI 1.28-1.41) were independently associated to flu vaccination. Heart and lung diseases were the morbidities associated with the higher odds of being vaccinated [aOR 1.97 (95%CI 1.86-2.09) and aOR 1.92 (95%CI 1.84-2.01), respectively]. Nursing home residents aged ≥ 65 years showed a lower odds of being vaccinated [aOR 0.39 (95%CI 0.28-0.54)]. Our data claims for an urgent public heath effort to fill the gap of missed vaccination opportunities reported in the past flu seasons.

## 1. Introduction

Influenza is an acute viral infection of the respiratory tract, whose clinical manifestations range from mild upper respiratory tract symptoms to life treating complications requiring hospitalization. It is estimated that every year flu can cause worldwide high morbidity and mortality epidemics with approximately 3 to 5 million cases and between 250,000 and 500,000 deaths [1]. In the northern hemisphere Influenza viruses circulating during fall and winter seasons cause epidemic outbreaks with an important burden of morbidity and a remarkable mortality rate. In Italy, the cost for the national health care system has been estimated an average of 1.4 US$ billion for each season between 1999 and 2008 [2].

The new pandemic caused by Severe Acute Respiratory Syndrome-Coronavirus-2 (SARS-CoV-2) has rapidly spread starting from China in late December 2019 and abruptly involving the north of Italy in late February 2020 [3]. SARS-CoV-2 infection is able to cause a severe respiratory syndrome with the requirement of intensive care assistance and eventually leading to death in a significant percentage of hospitalized patients [4, 5]. The dramatic impact of the SARS-CoV-2 pandemic on the Italian national healthcare system lead the national authorities to impose a national lock-down from March 9 to May 18, 2020. After a period of apparent control in summer months, current Italian epidemiological situation is evolving with a slight daily increment of the number of cases and a progressive increment of hospitalized patients [6]. Consequently, non-pharmacological interventions, such as the face mask combined with social distancing, are still enforced.

The new challenge in the management of the pandemic will appear in the 2020/2021 flu season, when the co-occurrence of influenza and flu-like illnesses may overcome the diagnostic capacity of our system with regard to the capability for molecularly testing all subjects with respiratory symptoms. Moreover, the clinical impact of subsequent or concomitant influenza and SARS-CoV-2 infections is still unknown, especially in the elderly populations and in patients with comorbidities.

To date, no effective treatment has been demonstrated to radically change the natural history of SARS-CoV-2 infection [7] and several vaccination strategies are still under development [8]; on the other hand, flu vaccination is able to decrease both the incidence rates of influenza and to moderate the severity of the disease in case of infection. Moreover, a remarkable decrease in morbidity for pneumonia, respiratory and cardiovascular complications and consequent risk of hospitalization and death has been consistently reported in the literature [9-12]. The reduced risk of hospitalization is particularly important considering the significant amount of hospital resources that are required for the management of COVID 19 patients which required hospitalization and intensive care assistance in a not negligible percentage [4, 5]. In the end, influenza vaccinations are not only safe, with few adverse events related to its administration, but also proven to be effective in children above 6 months of age, people with comorbid conditions and also in the elderly population [9, 10, 13].

Despite national and international recommendations for routine flu vaccination, vaccine uptake in Italy remains low in the general population, as well as in the high risk groups. Our aim was to assess the reported rate of flu vaccination in the season 2019/2020 in respondents to the nationwide online EPICOVID 19 survey [14] and to identify factors associated with the uptake of flu vaccination, in order to address potential gaps to be filled in the public health system.

## 2 Materials and Methods

### 2.1 Study design and setting

The study design was described elsewhere [14, 15] and the study was registered (ClinicalTrials.gov NCT04471701). In brief, EPICOVID19 is an Italian cross-sectional, web-based survey that was initiated in April 2020. A national convenience sample of volunteers aged 18 or older with access to internet and able to give an informed consent was assessed. The participants were recruited via social media (Facebook, Twitter, Instagram, Whatsapp), press releases, internet pages, local radio, TV stations, and institutional websites; the European Commission’s open-source official EUSurvey management tool was used to collect the data (https://epicovid19.itb.cnr.it/). The geographical coverage [14] such as the representativeness of the survey according to the Italian official demographic data [15] were described elsewhere.

### 2.2 Data collection and variables

For the purposes of this study, we analysed all the survey responses collected between 13^th^ April and 2^nd^ June 2020. The socio-demographic information included sex (male vs female, furtherly classified as pregnant or not), age (18-24, 25-29 up to 90+ and further categorized as <65 or ≥ 65 years of age), educational level (primary school or less = Low, middle or high school = Medium, and university degree or post-graduate = High), and occupational status (unemployed, employed, retired, student, and other). Work categories included in the national guidelines for flu vaccine were generated (Health professionals, armed forces, agricultural, forestry and fisher workers). Two ad hoc dummy variables were also created for categorizing residents in nursing homes (yes or not) and for respondents living with an “at risk cohabitant” according to the 2017-2019 Italian national vaccination plan (yes or not) [16]. Chronic conditions reported were lung diseases, heart diseases, metabolic disorders, renal diseases, oncological diseases, diseases of the immune system, liver diseases and pre-planned major surgical procedures. A new variable was created by summing the chronic conditions referred by participants (categorised as none, 1, 2 or ≥3 co-morbidities). Information about autonomy in carrying out daily activities, self-rated health (very bad, bad, adequate, good or very good) was also available. The participants were also asked if they had received an influenza vaccination during the previous autumn/winter season.

### 2.3 Study population and group definition

Out of the 207,341 respondents, the study population consisted of 198,822 participants who provided informed consent, and for whom the data were fully available for the analysis. For the aim of this study, participants were categorized as vaccinated or not vaccinated for flu during the 2019/2020 season.

### 2.4 Statistical analysis

Descriptive analyses were performed to show the numerical distributions, overall and by flu vaccine status, of all the variables that at a later stage were included as independent in the models applied for statistical inference. Continuous variables were expressed as mean [Standard Deviation (SD)] whereas categorical and ordinal ones as count and percentages. Flu vaccine rates were calculated for all classes of age and for all Italian regions, in this latter case also stratifying by sex (male vs female) and age (<65 vs ≥65 years). The association between the independent variables and the flu vaccine (0=not reported, 1=reported) was assessed by applying a multivariable binary logistic regression model. Adjusted Odds Ratios (aOR), together with their 95% Confidence Intervals (CI), were estimated for each independent variable, with p-level of statistical significance set at .05, two tails. The same regression model was used when restricting the population analysed to older people (≥65 years). All the statistical analyses were carried out using SPSS version 25 (IBM Corp) and STATA version 15.0 (StataCorp LP).

### 2.5 Ethics and consent to participate

The Ethics Committee of the National Institute for Infectious Diseases (Italian: Istituto Nazionale per le Malattie Infettive I.R.C.C.S. Lazzaro Spallanzani) approved the EPICOVID19 study protocol (Protocol No. 70, 12/4/2020). The participants were requested to give their informed consent when they first accessed the on-line platform. The study was carried out in accordance with the principles of the Declaration of Helsinki.

All data were handled and stored in accordance with the European Union General Data Protection Regulation (EU GDPR) 2016/679; data transfer included encrypting/decrypting and password protection.

## 3. Results

Between April and June 2020 198,822 participants filled the online EPICOVID 19 survey and 41,818 (21.0%) reported to have received a flu vaccination shot during the last influenza season (2019/2020). Respondents were prevalently females (59.7%) and aged between 18-64 years (85.9%) with a mean age of 48 (SD +/-14.7 years) (Table 1).

**Table 1:**
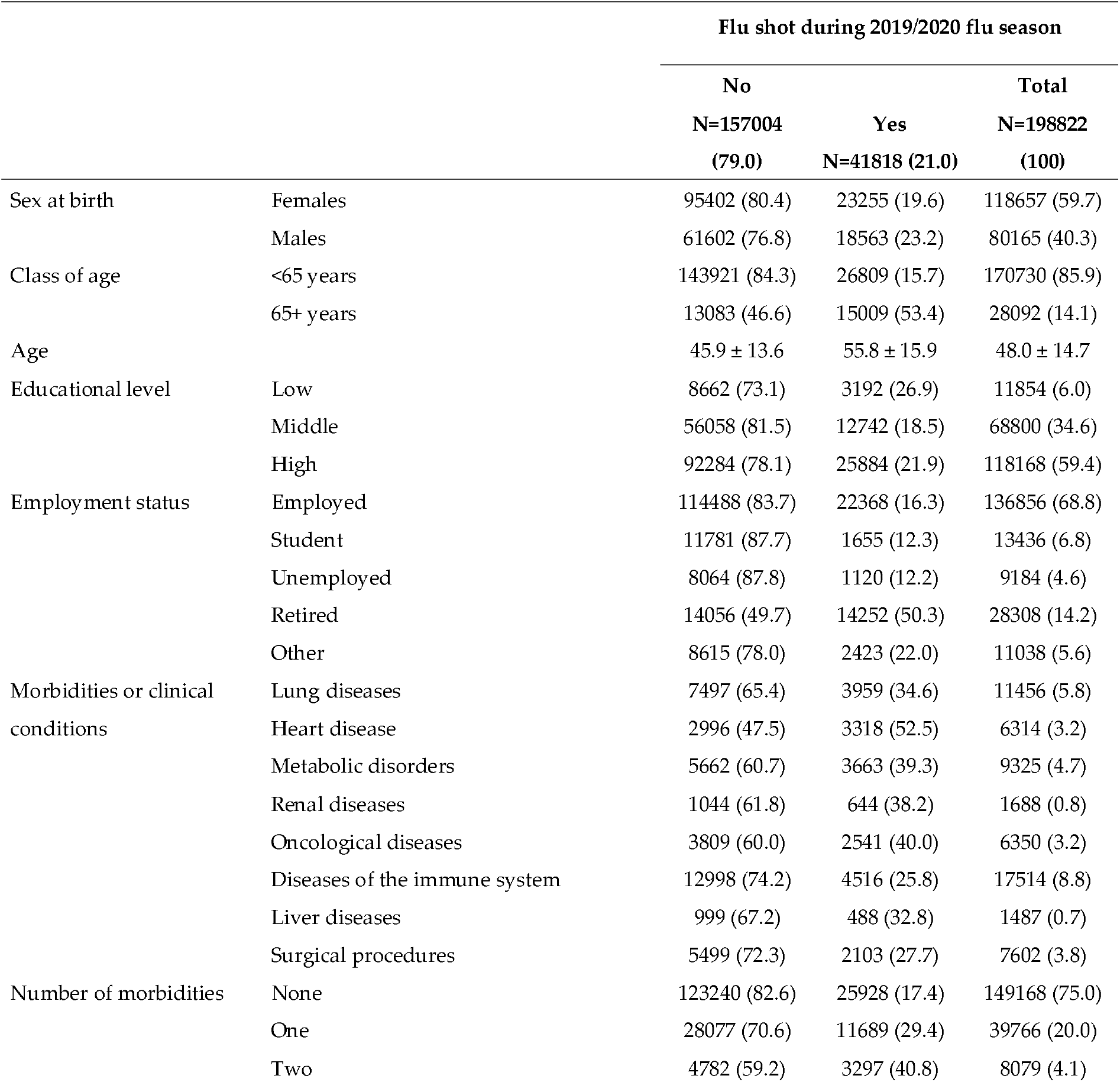
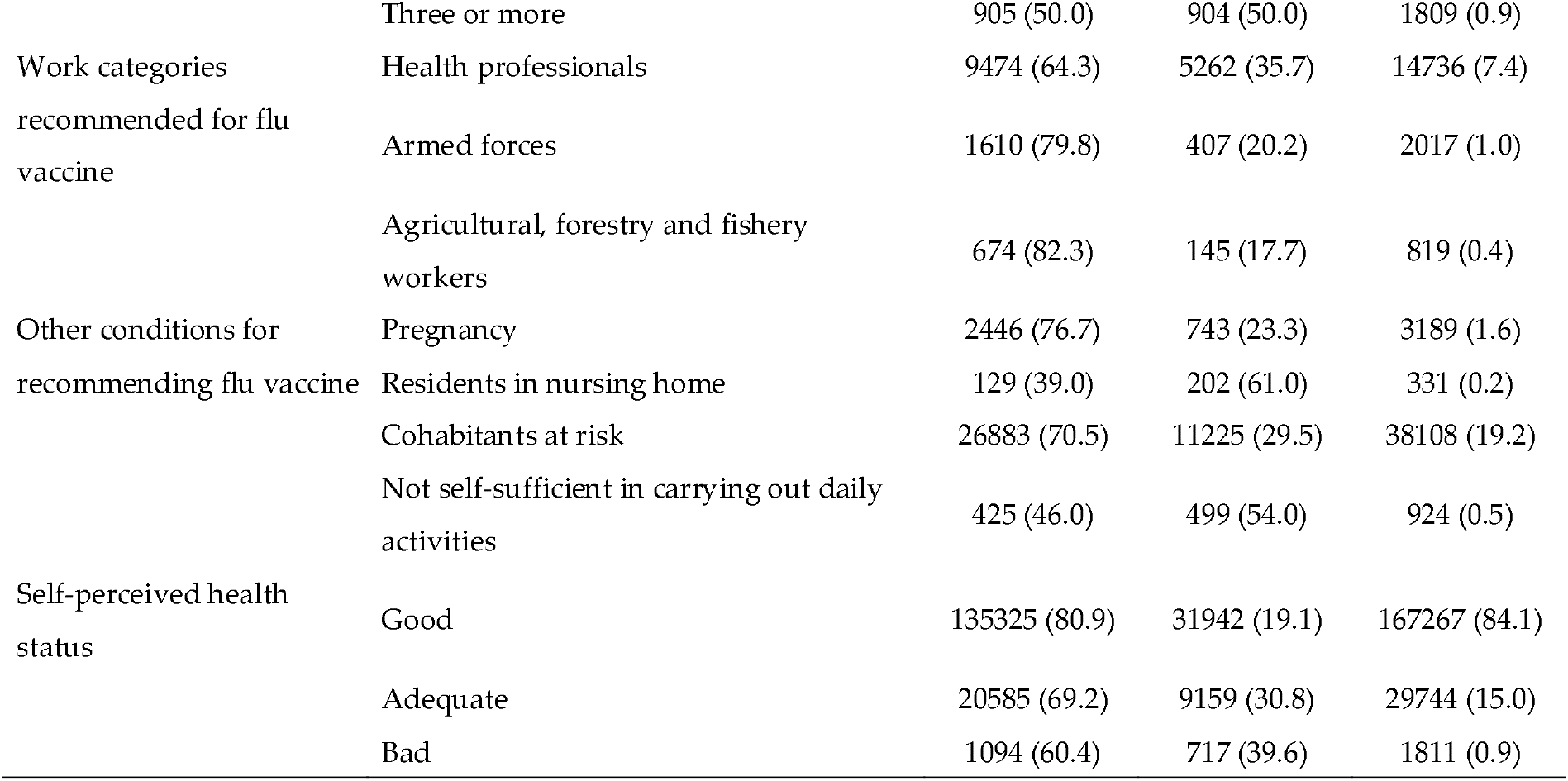
characteristics of study participants according to being or not vaccinated for influenza in the 2019/2020 flu season.

Vaccination coverage progressively increased with age, from 11% between 18 and 24 years of age and peaking to 72.9% between 85 and 89 years of age (Supplementary Table 1).

**Supplementary Table 1.** Vaccination coverage in participants according to the class of age.

| Class of age | Flu shot during 2019/2020 flu season |  |  |
| --- | --- | --- | --- |
|  | No | Yes | Total |
|  | N=157004 (79.0) | N=41818 (21.0) | N=198822 (100) |
| 18-24 | 8916 (89.0) | 1099 (11.0) | 10015 (5.0) |
| 25-29 | 12965 (87.6) | 1839 (12.4) | 14804 (7.4) |
| 30-34 | 15539 (87.1) | 2311 (12.9) | 17850 (9.0) |
| 35-39 | 17049 (85.6) | 2864 (14.4) | 19913 (10.0) |
| 40-44 | 17260 (85.0) | 3036 (15.0) | 20296 (10.2) |
| 45-49 | 19391 (86.4) | 3055 (13.6) | 22446 (11.3) |
| 50-54 | 19848 (86.0) | 3235 (14.0) | 23083 (11.6) |
| 55-59 | 18753 (81.7) | 4195 (18.3) | 22948 (11.5) |
| 60-64 | 14200 (73.3) | 5175 (26.7) | 19375 (9.7) |
| 65-69 | 7885 (55.3) | 6373 (44.7) | 14258 (7.2) |
| 70-74 | 3504 (41.1) | 5018 (58.9) | 8522 (4.3) |
| 75-79 | 1081 (35.1) | 2003 (64.9) | 3084 (1.6) |
| 80-84 | 379 (27.5) | 998 (72.5) | 1377 (0.7) |
| 85-89 | 136 (27.0) | 367 (73.0) | 503 (0.3) |
| 90+ | 98 (28.2) | 250 (71.8) | 348 (0.2) |

In particular, 15,009 (53.4%) subjects aged 65 years or older received a flu vaccination shot during the last influenza season. The comparison between vaccination covered per class of age and the official Italian from are reported in Supplementary Table 2.

**Supplementary Table 2.** Vaccination coverage per class of age according to the Italian official national data for the 2019/2020 flu season and in the EPICOVID19 respondents.

|  |  | Flu shot rates by class of age |  |
| --- | --- | --- | --- |
|  |  | National 2109-2020 | EPICOVID19 |
| Class of age | 18-44 years | 574,335/18,526,934 (3.1) | 11,149/82,878 (13.5) |
|  | 45-64 years | 1,749,887/18,227,994 (9.6) | 15,660/87,852 (17.8) |
|  | 18-64 years | 2,324,222/36,754,928 (6.3) | 26,809/170,730 (15.7) |
|  | 65+ years | 7,615,037/13,946,954 (54.6) | 15,009/28,092 (53.4) |
|  | 60-64 years | - | 5,175/19,375 (26.7) |

More than half of the respondents (59.4%) had high educational level, whereas 34.6% and 6% reported medium and high educational level. More than seventy-five percent of respondents (149,168; 75%) reported no diseases, whereas the remaining 25% reported at least one clinical condition representing *per se* a flu vaccine indication. Vaccination coverage progressively increased up to 50.0% in those reporting 3 or more comorbidities. Among participants reporting at least one condition, those with heart disease showed the higher vaccination coverage (52.5%) followed by those reporting oncological diseases (40%) and metabolic disorders (39%); whereas only 34.6% of those with lung diseases reported to be vaccinated for influenza. Among the professions with an indication for flu vaccination, health care workers were those with the higher coverage (35.7%).

Vaccination coverage ranged widely across Italian regions, from 18.2% in Abruzzo to 23.7% in Friuli-Venezia Giulia in the overall respondents (Figure 1A) and from 48.7% in Piemonte to 63.6% Friuli-Venezia Giulia in those aged ≥ 65 years (Figure 1B).

**Figure 1.**
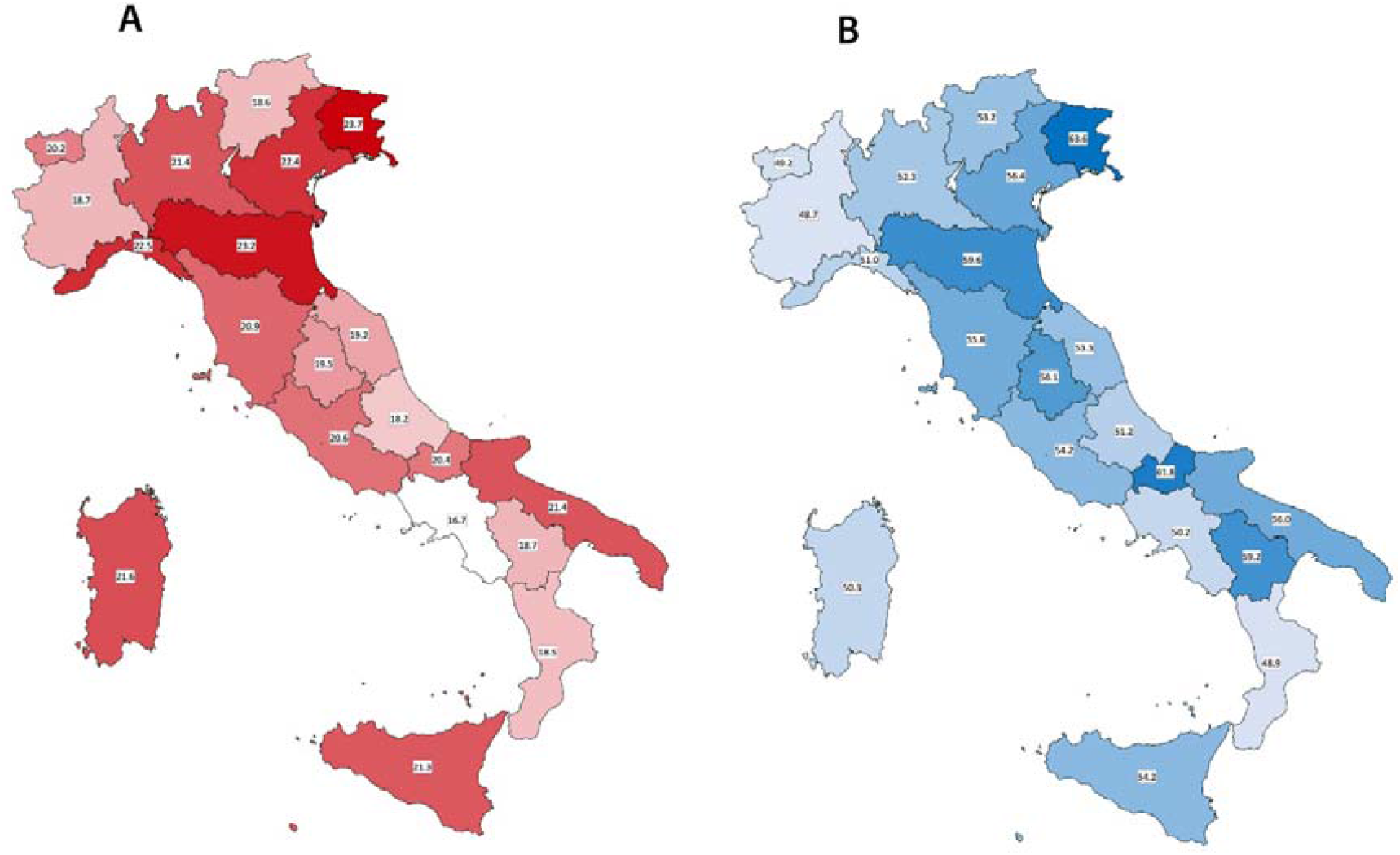
A, vaccination coverage according to the different Italian region in respondents aged ≥ 18 years. B, vaccination coverage according to the different Italian regions in respondents aged ≥ 65 years.

### 3.1 Multivariable model of factors associated to flu vaccination

Figure 2 shows the multivariable model for subjects aged 18 years or older.

**Figure 2.**
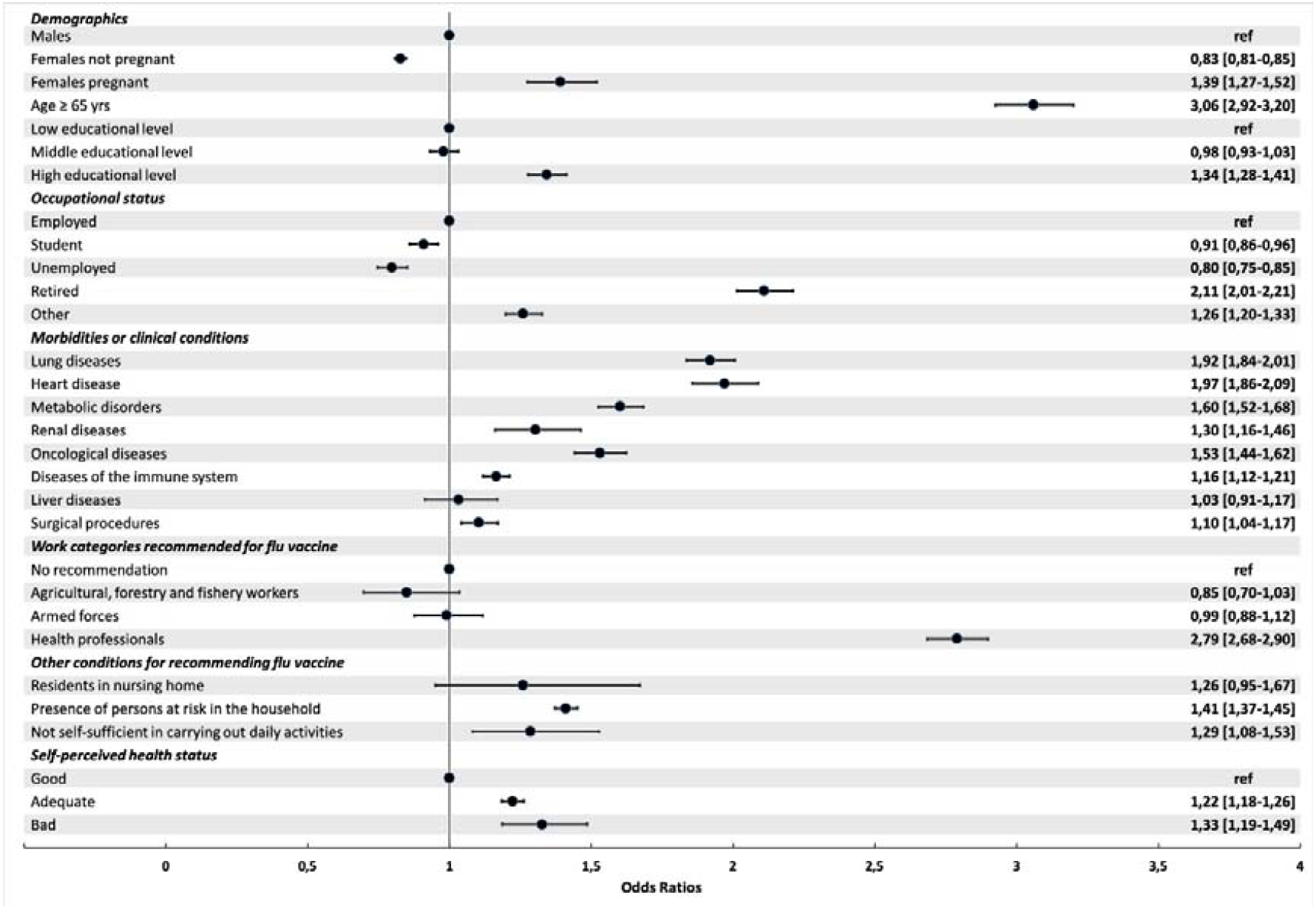
Multivariable model of factors associated to flu vaccination in the 2019/2020 season in respondents aged ≥ 18 years.

The multivariable model stratified by age (18-64 years and ≥65 years) are reported in Table 2. When considering the whole respondents, being 65 years or older (aOR 3.06, 95%CI 2.92-3.20) and having a high education level (aOR 1.34. 95%CI 1.28-1.41) were independently associated to flu vaccination. Conversely, those unemployed showed a reduced odds of being vaccinated (aOR 0.80, 95%CI 0.75-0.85).

**Table 2.**
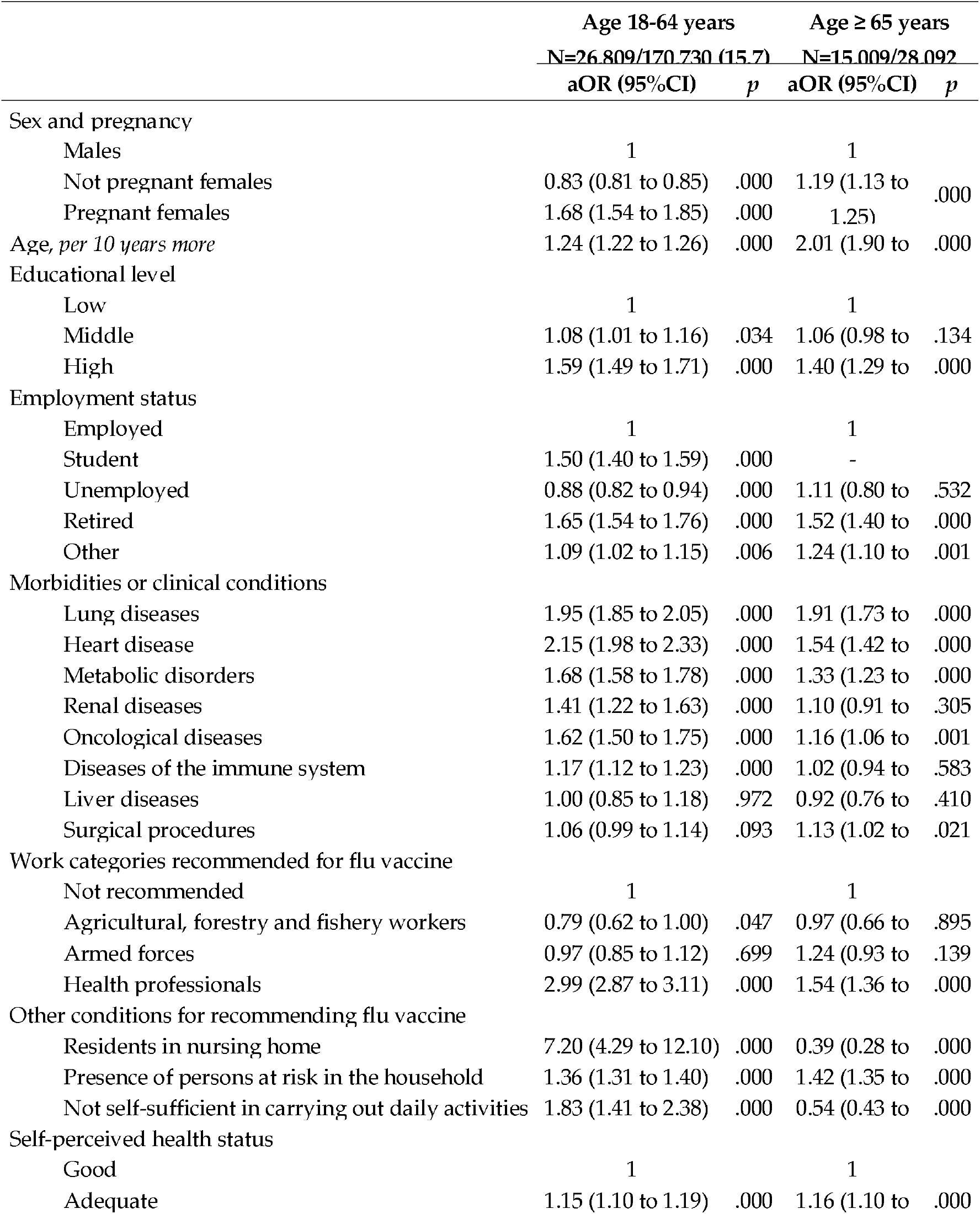
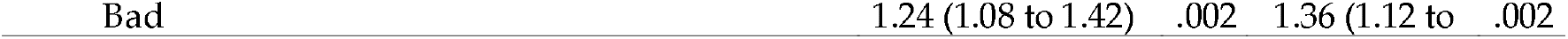
Multivariable binary logistic regression analysis on the probability of being or not vaccinated during the 2019/2020 flu season according to being 18-64 years old or ≥ 65 years old.

Among comorbidities, heart and lung diseases were conditions associated with higher odds of being vaccinated, both in subjects aged 18-64 [aOR 2.15 (1.98 to 2.33) and aOR 1.95 (1.85 to 2.05), respectively] and in those aged above 65 years [aOR 1.54 (95%CI 1.42-1.67) and aOR 1.91 (95%CI 1.73-2.12), respectively]. The odds of being vaccinated progressively increased with the number of reported morbidities, with a more pronounced effect in those aged below 65 years (Supplementary Table 3).

**Supplementary Table 3.** Multivariable binary logistic regression analysis on the probability of being or not vaccinated during the 2019/2020 flu season according to the number of reported morbidities.

| | Age 18-64 years | | Age $\geq 65$ years | |
| --- | --- | --- | --- | --- |
|  | N=26.809/170.730 (15.7) |  | N=15.009/28.092 (53.4) |  |
|  | aOR (95%CI) | p | aOR (95%CI) | p |
| Number of morbidities |  |  |  |  |
| None | 1 |  | 1 |  |
| One | 1.54 (1.49 to 1.59) | .000 | 1.43 (1.35 to 1.51) | .000 |
| Two | 2.18 (2.05 to 2.33) | .000 | 1.70 (1.55 to 1.86) | .000 |
| Three or more | 2.60 (2.26 to 2.99) | .000 | 1.66 (1.42 to 1.94) | .000 |

Health care workers showed a higher odds of being vaccinated [aOR 2.79 (95%CI 2.68-2.90)] when compared to work position without vaccine recommendation.

Living in a nursing home and being unable to perform daily activities were both associated with a lower odds of being vaccinated in those aged 65 years or older [aOR 0.39 (95%CI 0.28-0.54) and aOR 0.54 (95%CI 0.43-0.69), respectively].

### 4. Discussion

The EPICOVID 19 web based survey, which aimed to highlight different epidemiological and clinical aspects of COVID-19 epidemic in the first wave of the Italian epidemic, allowed the assessment of flu vaccination coverage in the 2019/2020 flu season in a large unselected sample of respondents. Moreover, we were able to identify predictors of flu vaccine uptake and highlight significant gaps in vaccination coverage in key populations such as in those residing in nursing homes and in those reporting specific medical conditions (i.e. liver diseases, kidney diseases and diseases of the immune system).

Our findings showed an overall vaccination rate of 21% and a rate of 53.4% in those aged 65 years or older, clearly below the target recommended coverage. The available National data obtained from a surveillance system collecting data from the general practitioners since 1990/2000 flu season, report a progressive increase in the vaccination coverage until 2009/2010 for the overall population (19.6 per 100 inhabitants) and until 2008/2009 for those aged 65 years or older (66.3 per 100 inhabitants). A progressive decline was subsequently observed for those aged 65 years or older until 2014/2015 (48.5 per 100 inhabitants) followed by a slight progressive increase in the subsequent years [17]. According to the official data of the Italian Ministry of Health of vaccination coverage in 2019/2020 flu season the overall vaccination coverage in the whole Italian population was 16.7 over 100 inhabitants and 54.6 over 100 inhabitants in those aged 65 years or older confirming the trend toward an increasing vaccination coverage [18]. Our data can not to be applied to the general estimates due to the absence in the survey of subjects aged 17 or younger. Whereas, the vaccination coverage observed in those aged 65 years or older appeared to be quite comparable to that reported by the official estimates (53.4% *vs* 54.6%, respectively). Although slightly increasing in recent years, the percentage observed in our study is lower than the target coverage of 75% in those aged 65 years or older. These data are particularly worrisome, because in the current fall/winter season in the northern hemisphere, with the concomitant co-circulation of both influenza and SARS-CoV-2 viruses, we will face a major challenge due to diagnostic and clinical problems in the presence of upper and lower respiratory tract symptoms [19]. Based on these concerns, for the 2020/2021 flu season the Italian ministry of Health recommends extending the vaccination coverage to the whole population, free of charge also for subject aged between 60 and 64 years [20]. If the free-of-charge administration of the influenza vaccine in this age-group will improve the uptake is to be demonstrated, but it will help to quantify the impact of the economical barrier to vaccination.

Recent findings support the need to increase the vaccination coverage, considering, on top of the well-known health and economic advantages, also the observed association between flu vaccination and reduction in SARS-CoV-2 nasopharyngeal swab positivity [15] and lower COVID-19 severity and mortality [21-23]. Therefore, the impact of vaccinations should be assessed not only based on their ability to prevent the related infections, but also on their broader impact on other communicable and not-communicable diseases occurring concurrently in the target population.

Our findings highlight a worrisome low vaccination coverage in subjects aged between 60-64 years and 65-69 years (26.7% and 44.7%, respectively). The low vaccination coverage in this group could be partially explained by a self-perception of wellbeing and low risk perception [24]. Nevertheless, these age groups are at high risk of COVID-19 complications and mortality, as observed in the Italian COVID-19 patients, therefore strategies to overcome their hesitancy towards vaccines and to increase vaccine accessibility and uptake are of paramount relevance [4, 25].

Another interesting finding observed is that people with higher education level seems to be better predisposed to vaccination when compared to the low socio-economic level. If on the one hand, low education level is a known general barrier to vaccines [26] on the other, our sample of generally higher educational level compared to the general population, reported overall a low uptake, meaning that multiple factors, beyond health literacy and access to health care services, need to be addressed. Moreover, the finding of a higher coverage among those with comorbidities suggest an attitude to consider the vaccination as a relevant intervention mainly for individuals at high risk, and not as a meaningful intervention for health promotion [27].

Our results showed that patients with heart and lung diseases seem to have a higher odds of being vaccinated when compared to those with other comorbidities, in particular kidney and liver disease. These findings raise particular concern, because it is clearly demonstrated that patients with chronic conditions, not only cardiovascular or respiratory diseases, are at higher risk of many vaccine-preventable diseases, including influenza, that increase the risk of serious complications, hospitalization, and death. Moreover, vaccine preventable diseases are more difficult to manage, because of increased risk of drug interactions and potential adverse effects [27].

In our study, flu vaccination resulted positively associated with being a health care worker, compared to other work-categories. Nevertheless, an extremely low percentage of health care workers respondents accounting for 35.7% received a flu shot in the last season. Although the percentage reported in our study is surprisingly higher compared to that reported in the literature in Italian health care workers, ranging from 14% [28] to 16% [29], it underlines the need for urgent measures to increase vaccination coverage in Italian health care workers. Vaccination of health care workers against influenza is a key component of infection control, and the rationale for it is based on the need to protect the health care workers and their high-risk patients in nursing homes, hospitals, and outpatient clinics. Moreover, in time of extreme demand, reducing the absenteeism due to influenza and its complications, contributes to the preservation of the essential health care services.

The finding that nursing home residents aged 65 years or older have a 61% higher odds of not being vaccinated compared to the rest of the population was unexpected and definitively worrisome. These subjects have high rates of underlying medical conditions and, therefore, are at higher risk of infectious disease outbreaks. As we have learned during the recent pandemic, about half of deaths due to COVID-19 were among nursing homes residents, underlying a critical health disparity that need to be addressed urgently. Increased vaccination rates and increased testing for preventable infectious disease of residents and staff, and increased access to personal protective measures and equipment are mandatory in all nursing homes, in order to protect this vulnerable subgroup of our population [30-32].

### 4.1 Strengths and limitations

The main strength of our research was the possibility to assess factors associated with flu vaccination in a large unselected group of respondents to an online survey. In fact, Italian estimates on past flu vaccination seasons reported aggregated data for the general population and for those aged 65 years or older but failed to address among the target group which are categories with significant barriers to achieve the pre-planned goal for flu vaccine coverage.

Our study has several limitations. First, the absence of subjects aged below 18 years makes impossible the comparison of our data with that of the Italian general population reported by the ministry of health. Second, the high percentage of high education and health care workers observed in our sample should be considered in the interpretation of the results for the possible impact on behaviours and opportunities toward a better health. Third, caution is warranted in the interpretation of flu vaccination coverage of nursing home residents due to the possibility of recalling bias. In the end, the sample cannot be considered fully representative of the Italian demographic curve and consequently our findings could be not completely generalizable to the Italian population.

## 5. Conclusions

Our data confirm a low vaccination coverage in the overall respondents to the online EPICOVID 19 survey and in particular, far below the national recommendations in target populations, such as those 65 years or older, in subjects at high risk due to chronic conditions and in health care workers. In the perspective of the next fall winter season, with the probable co-circulation of influenza and SARS-CoV-2 in the norther hemisphere, the population will remain vulnerable to concurrent epidemics. The morbidity and mortality impact will be directly related to the strength of the public health response, which must be based on effective infection prevention tools currently available: the non-pharmacologic interventions (i.e. social distance and face masks) and the widespread implementation of influenza vaccination to fill the gap of missed vaccination opportunities reported in the past flu seasons.

## Data Availability

The data will be available upon reasonable request.

## Supplementary Materials

The following are available online at www.mdpi.com/xxx/s1,

## Author Contributions

Conceptualization, A.G, M.G., S.M., F.P., F.A.; Formal analysis, F.P., F.A; Methodology, A.G, M.G., S.M., F.P. and F.A.; Project administration, F.A.; Supervision, M.G. and F.A.; Writing—original draft, A.G., M.G., F.P., S.M. and F.A.; Writing—review & editing, S.M, M.N., C.T., R. A.I, G.P., G.B, M.D.B, A.S., M.T., C.M., M.A., F.P., F.A. The corresponding author, A.G., attests that all listed authors meet authorship criteria and that no others meeting the criteria have been omitted. All authors have read and agreed to the published version of the manuscript.

## Acknowledgments

The EPICOVID19 Working Group: Fulvio Adorni (National Research Council, Institute of Biomedical Technologies, Via Fratelli Cervi 93, 20090 Segrate (MI), Italy); Federica Prinelli (National Research Council, Institute of Biomedical Technologies, Via Fratelli Cervi 93, 20090 Segrate (MI), Italy); Fabrizio Bianchi (National Research Council, Institute of Clinical Physiology, Via G. Moruzzi 1, 56124 Pisa (PI), Italy); Andrea Giacomelli (Infectious Diseases Unit, Department of Biomedical and Clinical Sciences L.Sacco, Università di Milano, ASST Fatebenefratelli Sacco, Milan, Italy Via G.B. Grassi 74, 20157 Milano, Italy); Gabriele Pagani (Infectious Diseases Unit, Department of Biomedical and Clinical Sciences L.Sacco, Università di Milano, ASST Fatebenefratelli Sacco, Milan, Italy Via G.B. Grassi 74, 20157 Milano, Italy); Dario Bernacchia (Infectious Diseases Unit, Department of Biomedical and Clinical Sciences L.Sacco, Università di Milano, ASST Fatebenefratelli Sacco, Milan, Italy Via G.B. Grassi 74, 20157 Milano, Italy); Stefano Rusconi (Infectious Diseases Unit, Department of Biomedical and Clinical Sciences L.Sacco, Università di Milano, ASST Fatebenefratelli Sacco, Milan, Italy Via G.B. Grassi 74, 20157 Milano, Italy); Stefania Maggi (National Research Council, Neuroscience Institute, Aging branch, Via Vincenzo Maria Gallucci 16, 35128 Padova, Italy); Caterina Trevisan (University of Padova, Department of Medicine (DIMED) – Geriatric Unit, Via Giustiniani 2, 35128 Padova (PD), Italy; National Research Council, Neuroscience Institute, Via Giustiniani 2, 35128 Padova (PD), Italy); Marianna Noale (National Research Council, Neuroscience Institute, Aging branch, Via Vincenzo Maria Gallucci 16, 35128 Padova, Italy); Sabrina Molinaro (National Research Council, Institute of Clinical Physiology, Via G. Moruzzi 1, 56124 Pisa (PI), Italy); Luca Bastiani (National Research Council, Institute of Clinical Physiology, Via G. Moruzzi 1, 56124 Pisa (PI), Italy); Loredana Fortunato (National Research Council, Institute of Clinical Physiology, Via G. Moruzzi 1, 56124 Pisa (PI), Italy); Nithiya Jesuthasan (National Research Council, Institute of Biomedical Technologies, Via Fratelli Cervi 93, 20090 Segrate (MI), Italy); Aleksandra Sojic (National Research Council, Institute of Biomedical Technologies, Via Fratelli Cervi 93, 20090 Segrate (MI), Italy); Carla Pettenati (National Research Council, Institute of Biomedical Technologies, Via Fratelli Cervi 93, 20090 Segrate (MI), Italy); Marcello Tavio (Division of Infectious Diseases, Azienda Ospedaliero Universitaria Ospedali Riuniti, Ancona, Italy); Massimo Andreoni (Infectious Diseases Clinic, Department of System Medicine, Tor Vergata University of Rome, 00133 Rome, Italy); Claudio Mastroianni (Public Health and Infectious Disease Department, “Sapienza” University, Rome, Italy); Raffaele Antonelli Incalzi (Unit of Geriatrics, Department of Medicine, Biomedical Campus of Rome, via Alvaro del Portillo, 21, 00128 Rome, Italy); Massimo Galli (Infectious Diseases Unit, Department of Biomedical and Clinical Sciences L.Sacco, Università di Milano, ASST Fatebenefratelli Sacco, Milan, Italy Via G.B. Grassi 74, 20157 Milano, Italy).

